# Geospatial Variability in Excess Death Rates during the COVID-19 Pandemic in Mexico: Examining Socio Demographic and Population Health Characteristics

**DOI:** 10.1101/2021.08.11.21261930

**Authors:** Sushma Dahal, Ruiyan Luo, Monica H. Swahn, Gerardo Chowell

**Affiliations:** Department of Population Health Sciences, School of Public Health, Georgia State University, Atlanta, GA, USA; Wellstar College of Health and Human Services, Kenessaw State University, Kenessaw, GA, USA

**Author notes:** Corresponding author: (SD).

**Keywords:** excess mortality, COVID-19 pandemic, Mexico, states, socio-demographic factors, spatial variation

## Abstract

**Background:** Mexico has suffered one of the highest COVID-19 mortality rates in the world. In this study we examined how socio demographic and population health characteristics shape the geospatial variability in excess mortality patterns during the COVID-19 pandemic in Mexico.

**Methods:** Weekly all-cause mortality time series for all 32 Mexican states, from January 4, 2015 to April 10, 2021, were analyzed to estimate the excess mortality rates using Serfling regression models. The association between socio-demographic, health indicators and excess mortality rates were determined using multiple linear regression analyses. Finally, we used functional data analysis to characterize clusters of states with distinct mortality growth rate curves.

**Results:** The overall all-cause excess deaths rate during the COVID-19 pandemic in Mexico until April 10, 2021 was estimated at 39.66 per 10 000 population. The lowest excess death rates were observed in southeastern states including Chiapas (12.72), Oaxaca (13.42) and Quintana Roo (19.41) whereas Mexico City had the highest excess death rate (106.17), followed by Tlaxcala (51.99) and Morelos (45.90). We found a positive association of excess mortality rates with aging index (P value<.0001), marginalization index (P value<.0001), and average household size (P value=0.0003) in the final adjusted model (Model R^2^=76%). We identified four distinct clusters with qualitatively similar excess mortality curves.

**Conclusion:** Central states exhibited the highest excess mortality rates whereas the distribution of aging index, marginalization index, and average household size explained the variability in excess mortality rates across Mexico. Our findings can help tailor interventions to mitigate the mortality impact of the pandemic.

**Key message:**

- This study quantified and examined spatial patterns of excess mortality across states of Mexico, with lower rates of excess mortality in southeastern states and higher rates in central states.
- Mexico City recorded 1 in 5 of all excess deaths in reported in Mexico, which accounted for 19% of total excess deaths across the country.
- Findinds indicate that aging index, marginalization index, and average household size played a significant role on excess death rates across Mexican states during the COVID-19 pandemic.
- Four distinct clusters characterized the excess mortality curves across Mexican states.

## Introduction

Monitoring all-cause excess mortality, above an expected level of total deaths, as a pandemic unfolds is one of the key ways to evaluate its mortality impact (1). All-cause excess mortality estimates include deaths that are directly or indirectly attributed to the pandemic (2, 3). While the excess deaths can be directly attributed to COVID-19, other excess deaths attributed to the pandemic such as those that could be related to denied or delayed care for acute emergencies (4) (5) or other chronic conditions (6), the disruption of routine health care services in an overburdened health care system(7), unaddressed mental health concerns including suicide and self-harm (8, 9), and drug overdoses (10). Detailed analyses of excess mortality can help determine where the mortality impact of the pandemic has been most significant.

Mexico is one of the countries in Latin America that is bearing the brunt of COVID-19 pandemic with the fourth-highest number of COVID-19 deaths in the world, after the USA, Brazil, and India, as of late June 2021 (11). In fact, Mexico has reported a total of 2 487 747 (1.38% of global cases) confirmed cases of COVID-19, including 231 847 deaths (5.96% of global deaths), as of June 25, 2021 (12). A previous study reported a high all-cause excess death rate of 26.10 per 10 000 population in Mexico in 2020, with COVID-19 deaths accounting for only 38.64% of the estimated excess deaths (13). While this relatively low proportion of COVID-19 deaths out of all excess deaths could be the result of low testing rates, misclassification of COVID-19 deaths, and delays in reporting COVID-19 deaths (14), a substantial number of deaths during the pandemic could be due to the indirect causes (3) and need to be examined in more depth.

The distribution of indirect causes of deaths depends on several factors such as sociodemographic characteristics, population health and the selection, timing and intensity of any public health interventions, in addition to the efficiency and reach of the health and social care system (15). In Mexico, pandemic control measures have varied widely (16). Therefore, a more detailed understanding of the mortality burden of the pandemic can be obtained by quantifying spatial heterogeneity in excess deaths at a state level and by examining the influence of underlying sociodemographic, economic, and health system related factors. In this study, we pose the question whether in a country such as Mexico, with very high covid-19 mortality, potential spatial variability in the excess deaths can be explained by underlying sociodemographic and population health indicators. To answer that research question, we first estimated the all-cause excess deaths during the COVID-19 pandemic in Mexico comprising 31 states and Mexico City. Next, we evaluated the potential associations between different socio-demographic factors and excess mortality patterns at the state level in Mexico. Furthermore, we also conducted a cluster analysis to characterize the shapes of the excess mortality curves into different groups that describe the potential geospatial variability in excess mortality. Analyses such as these are critically important for understanding excess mortality and for guiding intervention strategies.

## Methods

### Data

We obtained weekly all-cause death counts updated on May 25, 2021 for Mexico at the state level and for Mexico City, based on epidemiological weeks from January 2020 until April 10, 2021 and for the preceding 5□years (2015–2019) to establish a baseline mortality level(17). We accessed publicly available weekly mortality data from the National Institute of Statistics and Geography (INEGI) for the years from 2015 to 2018, and data from National Population Registry (RENAPO) for the years 2020 and 2021(17). For the year 2019, we chose either INEGI or RENAPO as the data source, based on the value of the last week of 2018 and the first week of 2019 for each state. We obtained the national and state-level population size estimates from the National Population Council (CONAPO) of Mexico(18). Mortality data was not available for the state of Tlaxcala for the last six weeks of the study period. For this reason, this state was excluded from our regression and functional cluster analyses.

For each state, including Mexico City, we obtained data on six variables: population density (2020), aging index (2020), average household size (2020), marginalization index (2020), rate of new cases of depression per 100 000 population (2019), and public spending on health as percent of GDP (2019). Data on population density, aging index, average household size, and rate of new case of depression were obtained from INEGI (19), data on public spending on health was obtained from the subsystem of health accounts at the federal and state level (SICUENTAS)(20), and the data on the marginalization index was available from CONAPO(21). Summary statistics of these variables from 31 states and Mexico city are provided in Table 1.

**Table 1.** Descriptive statistics for six study variables included in the multiple regression analysis of Excess Mortality in Mexico (*n*=32)

| Variable | Minimum | Mean (SD) | Median (IQR) | Maximum |
| --- | --- | --- | --- | --- |
| Population density (2020)<br>(habitants per km<sup>2</sup>) | 10.80 | 309.68 (1078.69) | 67.15 (127.20) | 6163.30 |
| Aging index (2020) | 28.70 | 46.41 (10.44) | 45.45 (7.40) | 90.20 |
| Average household size (2020) | 3.6 | 3.91 (0.18) | 3.90 (0.20) | 4.4 |
| Marginalization index (2020) | 11.32 | 18.89 (2.73) | 19.43 (3.23) | 23.01 |
| Rate of new case of depression<br>per 100,000 population (2019) | 22.06 | 114.88 (76.05) | 92.14 (67.37) | 348.17 |
| Public spending on health as a percent of GDP (2019) | 0.94 | 3.12 (1.02) | 2.96 (1.32) | 5.81 |

### Pandemic period and excess deaths

For both the national data and the data for each state, we separately estimated the baseline mortality level by fitting cyclical Serfling regression models to all-cause deaths in the non-COVID-19 period, after excluding data from March 2020 to April 2021 by employing established methodology (2, 22-25). Details on the model equation that was used can be found in ref (13). After establishing a weekly baseline and the corresponding 95% CI at the national level, we defined the periods of COVID-19 pandemic as the weeks in 2020 and 2021 where the observed all-cause mortality rate at the national level in Mexico exceeded the upper 95% confidence limit of the national baseline mortality rate. The excess mortality rate was estimated at the state level and for Mexico City for the same defined period of COVID-19 pandemic. Excess all-cause mortality rate was estimated as the difference between the observed and model adjusted baseline mortality rates for each week constituting the pandemic period. The overall pandemic excess mortality in 2020 and 2021 was calculated by summing the excess death rates across the pandemic weeks in the given year (13, 22, 24). Negative excess mortality estimates were replaced by zeros in our analyses to account for underreporting due to reporting delay (3, 26).

### Multiple regression analysis

After estimating the total excess mortality rate for each state, we explored the association between the total excess mortality rate and the predictor variables. Because the population density and rate of new case of depression distributions were skewed, we transformed these variables to log base 10. Since we identified Mexico City as a potential influential point, we performed sensitivity analysis by comparing the results of different models, including and excluding Mexico City. Since there was no significant change in the statistical inference of the parameters, we included Mexico City in the multiple regression analysis.

### Cluster analysis

We followed the analytic methods described in (27) to pre-process the weekly cumulative all-cause excess deaths for 30 states and Mexico city (excluding Tlaxcala, refer to study setting for details). Then, we analyzed the shapes of the excess all-cause death rate curves to compare, cluster, and summarize growth rates.

We employed the following steps to smooth and normalize the weekly all-cause excess death data:

a. Smoothing: Cumulative excess deaths curves were smoothed using smooth function in Matlab which uses a moving average filter over a 10-week span.
b. Time differencing: If *f*_*i*_(*t*) denotes the given cumulative number of excess deaths for state i on week t, then per week growth rate at time t is given by *g*_*i*_(*t*) = *f*_*i*_(*t*) − *f*_*i*_(*t* − 1).
c. Re-scaling: We rescaled each curve by dividing each *g*_*i*_(*t*) by the total excess number of deaths for a given state i, which is equivalent to computing *h*_*i*_(*t*) = *g*_*i*_(*t*) / *r*_*i*_ where 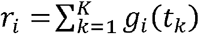 and K is the number of weeks in the period.
d. Smoothing: We then smoothed the normalized curves over a 5-weeks span, using the smooth function in Matlab.

To identify the clusters by comparing the curves, we used a simple metric. For any two rate curves, *h*_*i*_ and *h*_*j*_, we compute the norm ∥h_i_ −h_j_∥, where the double bars denote the *L*^*2*^ norm of the difference function, i.e., 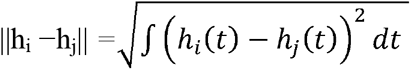, which is approximated by 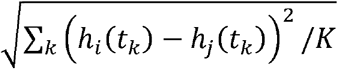, where K is the number of weeks in the period.

To perform clustering of thirty-one curves into smaller groups, we applied the dendrogram function in Matlab using the “ward” linkage as explained in ref. (27). The number of clusters was decided empirically by inspecting the overall clustering results. After clustering the states into different groups, we derived the average curve for each cluster using a time wrapping algorithm (27, 28).

## Results

Observed death rate was greater than the upper 95% confidence interval of the baseline in 52 weeks starting from week of April 12-18, 2020, out of total 58 weeks from March 1, 2020, to April 10, 2021 (Figure 1). The excess mortality rate first peaked during the week of July 12-18, 2020, with the excess death rate of 1.04 per 10 000 population, then declined slightly for a few weeks and then increased again from the week of December 20-26, 2020, and reached a peak with an all-cause excess death rate of 1.99 per 10 000 population on the week of January 17-23, 2021. The excess death rate remained below 0.5 since the week of 28 February 2021 until the end of the study period.

**Figure 1.**
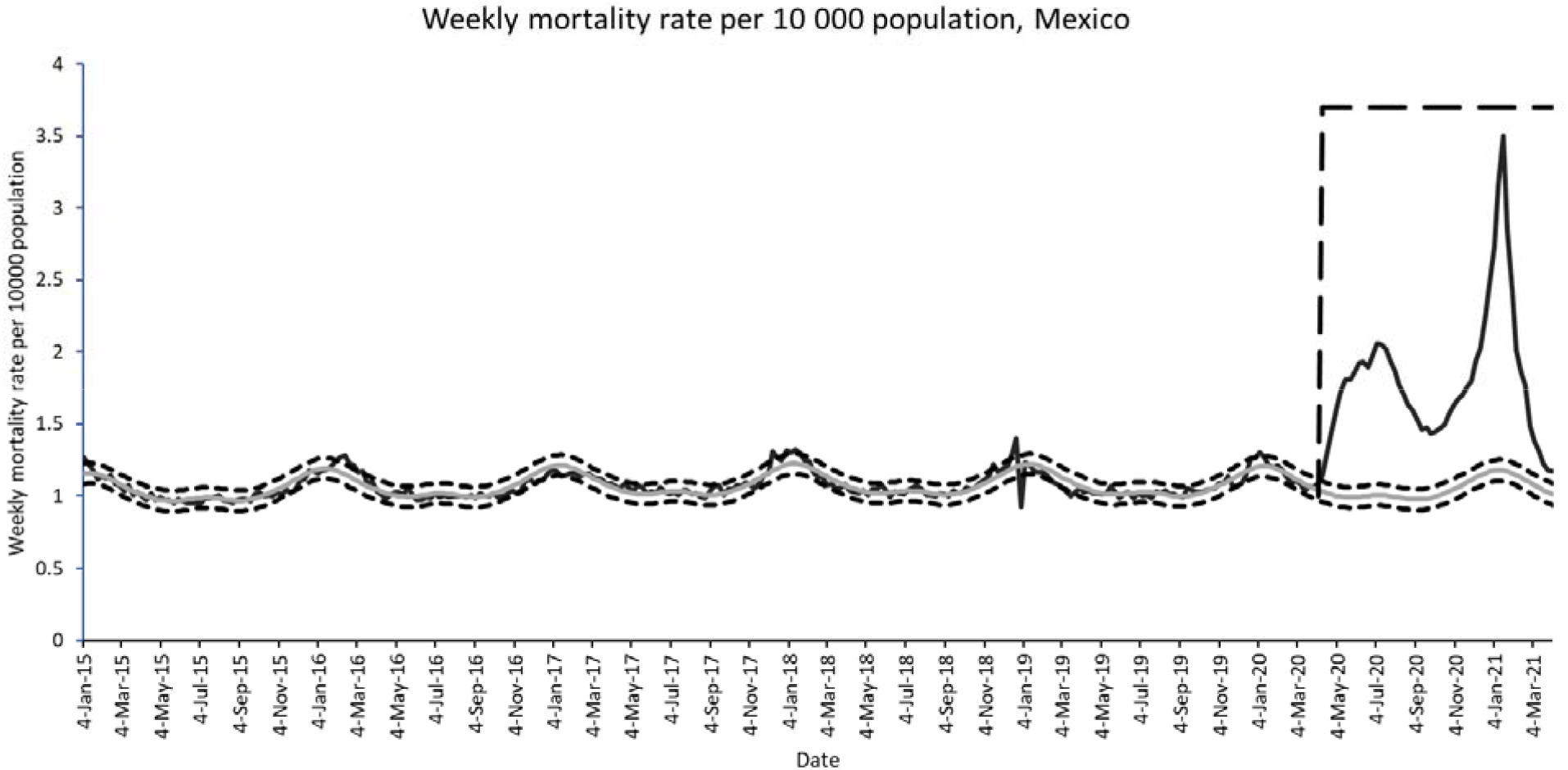
Mortality rate per 10 000 population, Mexico, January 2015–March 2021. The black curve is the observed weekly death rate. The grey curve is the predicted baseline death rate. Square dotted curves indicate the upper and lower 95% confidence intervals of the baseline death rate. The long dashed line indicates the COVID-19 pandemic period.

Table 1 shows the total all-cause excess death rate for the national level, Mexico City, and 31 states of Mexico. The map showing the estimates at the state level is given in Figure 2. While the total excess death rate in Mexico was at 39.66 per 10 000 population, equivalent to a total of ∼508 289 excess deaths, the excess mortality rate in Mexico City was the highest and estimated at 106.17 per 10 000 population (∼95 690 total number of excess deaths). Among 31 states, Tlaxcala (51.99), Morelos (45.90), Puebla (45.12), and Mexico (44.43) were among the states with the highest excess mortality rates. The states with the lowest death rates included Chiapas (12.72), Oaxaca (13.42), Quintana Roo (19.41), and Yucatan (21.11) (Table 2). Only one state, Chiapas, had no excess deaths in 2021. COVID-19 accounted for only 42.16% of total exces deaths at the national level ranging from 20.97% in Chiapas to 76.05% in Quintana Roo.

**Table 2.** Estimates for all cause excess mortality rate by state per 10 000 population during COVID-19 pandemic in Mexico, March 1, 2020-April 10, 2021

| State/Region | All cause excess death rate per 10 000 population (March 1, 2020-April 10, 2021) | All cause excess death rate per 10 000 population in 2020 (Includes weeks starting from March 1, 2020, to December 27, 2020) | All cause excess death rate per 10 000 population in 2021 (Includes week starting on January 3, 2021, to April 4, 2021) | Total number of all cause excess deaths | COVID-19 deaths (Percentage of all cause excess deaths) |
| --- | --- | --- | --- | --- | --- |
| National | 39.66 | 27.25 | 12.41 | 508 288.78 | 214 298<br>(42.16) |
| Aguascalientes | 29.37 | 21.61 | 7.76 | 4228.25 | 2300 (54.39) |
| Baja California Sur | 26.13 | 16.36 | 9.77 | 2118.75 | 1326 (62.58) |
| Baja California | 38.55 | 31.78 | 6.77 | 14 051.54 | 8048 (57.27) |
| Campeche | 22.25 | 21.02 | 1.22 | 2228.05 | 1183 (53.09) |
| Chihuahua | 27.76 | 25.37 | 2.39 | 10 561.33 | 6467 (61.23) |
| Chiapas | 12.72 | 12.72 | 0 | 7287.73 | 1528 (20.97) |
| Mexico City | 106.17 | 62.93 | 43.24 | 95 689.73 | 32 166 (33.61) |
| Coahuila | 38.33 | 30.88 | 7.45 | 12 369.31 | 6189 (50.03) |
| Colima | 28.32 | 19.23 | 9.09 | 2234.78 | 1158 (51.82) |
| Durango | 27.48 | 22.94 | 4.53 | 5142.23 | 2372 (46.13) |
| Guerrero | 29.96 | 21.51 | 8.45 | 10 966.69 | 4231 (38.58) |
| Guanajuato | 36.54 | 20.44 | 16.09 | 22 842.82 | 10 568 (46.26) |
| Hidalgo | 36.39 | 22.86 | 13.52 | 11 277.96 | 5995 (53.16) |
| Jalisco | 32.92 | 18.66 | 14.26 | 27 799.37 | 11 759 (42.30) |
| Mexico | 44.43 | 28.67 | 15.76 | 77 705.35 | 33 571 (43.20) |
| Michoacan | 31.12 | 16.69 | 14.43 | 15 063.24 | 5492 (36.46) |
| Morelos | 45.90 | 24.16 | 21.75 | 9428.76 | 3052 (32.37) |
| Nayarit | 23.64 | 15.08 | 8.55 | 3060.99 | 1766 (57.69) |
| Nuevo Leon | 36.80 | 24.98 | 11.82 | 20 735.12 | 9305 (44.87) |
| Oaxaca | 13.42 | 10.55 | 2.87 | 5565.51 | 3494 (62.78) |
| Puebla | 45.12 | 36.50 | 8.62 | 29 849.45 | 11 142 (37.33) |
| Queretaro | 35.86 | 18.79 | 17.06 | 8241.92 | 4063 (49.29) |
| Quintana Roo | 19.41 | 16.95 | 2.46 | 3354.42 | 2551 (76.05) |
| San Luis Potosi | 32.26 | 22.29 | 9.97 | 9266.51 | 5190 (56.01) |
| Sinaloa | 31.53 | 24.97 | 6.56 | 9969.06 | 5927 (59.45) |
| Sonora | 32.22 | 27.11 | 5.11 | 9924.27 | 6485 (65.34) |
| Tabasco | 22.49 | 20.34 | 2.15 | 5791.32 | 3994 (68.96) |
| Tamaulipas | 30.33 | 25.38 | 4.95 | 11 086.40 | 4800 (43.29) |
| Tlaxcala | 51.99 | 34.93 | 17.06 | 7200.90 | 2367 (32.87) |
| Veracruz | 22.35 | 17.46 | 4.89 | 19 111.79 | 9524 (49.83) |
| Yucatan | 21.11 | 16.49 | 4.62 | 4781.42 | 3563 (74.52) |
| Zacatecas | 43.22 | 29.81 | 13.41 | 7217.52 | 2721 (37.69) |

**Figure 2.**
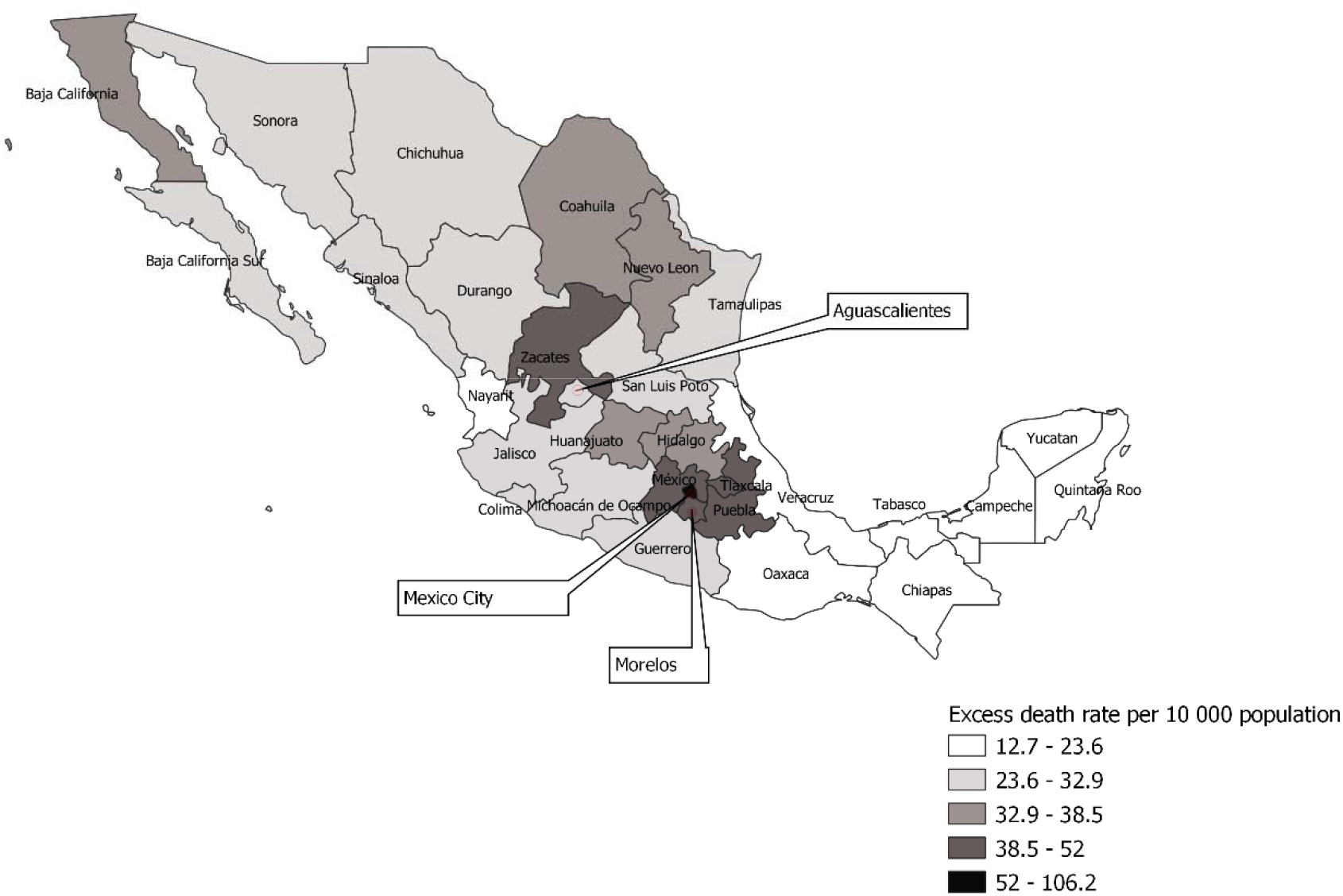
Map showing the excess death rate per 10 000 population at state level in Mexico.

Table 3 shows the results of fitting a taxonomy of multiple regression models of excess mortality rate at the state level in Mexico. Since R^2^ was slightly different for Models 4, 5, 6, and 7, we performed a multiple partial F-test to select the final model. We failed to find a significant contribution of adding population density, depression rate, and public expenditure on health on predicting excess mortality rate after accounting for the contribution of aging index, marginalization index, and average household size (F-value_3,24_=0.39, P-value=0.7631). Therefore we selected Model 4 as the final model. Our final model was able to explain 76% of the observed variance in the excess mortality rate (Coefficient of determination (R^2^)=0.76).

**Table 3.** Results of fitting a taxonomy of multiple regression models of excess mortality rate at the state level in Mexico (*n*=31)

|  | Parameter estimate<br>(*se*) |  |  |  |  |  |  |
| --- | --- | --- | --- | --- | --- | --- | --- |
|  | Model 1 | Model 2 | Model 3 | **Model 4** | Model 5 | Model 6 | Model 7 |
| intercept | - 19.21*<br>(9.03) | -18.18<br>(18.09) | -<br>47.03**<br>(13.61) | **-250.80**<br>***<br>**(50.67)** | -<br>215.08**<br>(63.11) | -<br>230.65**<br>(69.68) | -<br>229.07**<br>(72.97) |
| Aging index | 1.12***<br>(0.19) |  | 1.01***<br>(0.18) | **1.12\*\*\***<br>**(0.15)** | 0.99***<br>(0.20) | 0.94***<br>(0.22) | 0.94***<br>(0.22) |
| Marginalization index |  | 2.72**<br>(0.95) | 1.76*<br>(0.68) | **3.30\*\*\***<br>**(0.66)** | 2.98***<br>(0.74) | 2.94***<br>(0.76) | 2.87**<br>(1.02) |
| Average household size |  |  |  | **43.47\*\*\***<br>**(10.56)** | 35.64*<br>(13.39) | 37.95*<br>(14.18) | 37.95*<br>(14.47) |
| log10popdensity |  |  |  |  | 3.79<br>(3.98) | 4.38<br>(4.17) | 4.49<br>(4.41) |
| log10depression_rate |  |  |  |  |  | 4.20<br>(7.47) | 4.24<br>(7.63) |
| Public expenditure on health<br>as a percent of GDP |  |  |  |  |  |  | -0.22<br>(2.29) |
| Root MSE | 10.97 | 14.41 | 10.04 | **8.01** | 8.02 | 8.13 | 8.30 |
| $R^2$ | .55 | 0.22 | .63 | **0.76** | 0.78 | 0.78 | 0.79 |
| Model $F$-test | 35.18*** | 8.18** | 24.31*** | **31.09\*\*\*** | 23.46*** | 18.34*** | 14.68*** |
| ($df_1$, $df_2$) | (1, 29) | (1, 29) | (2, 28) | **(3, 27)** | (4, 26) | (5, 25) | (6, 24) |

\* $P < 0.05$ , \*\* $P < 0.01$ , \*\*\* $P < 0.001$

As shown in table 4, we found a positive association of excess mortality rate with aging index, marginalization index, and average household size in the adjusted model at 0.05 level of significance.

**Table 4.**
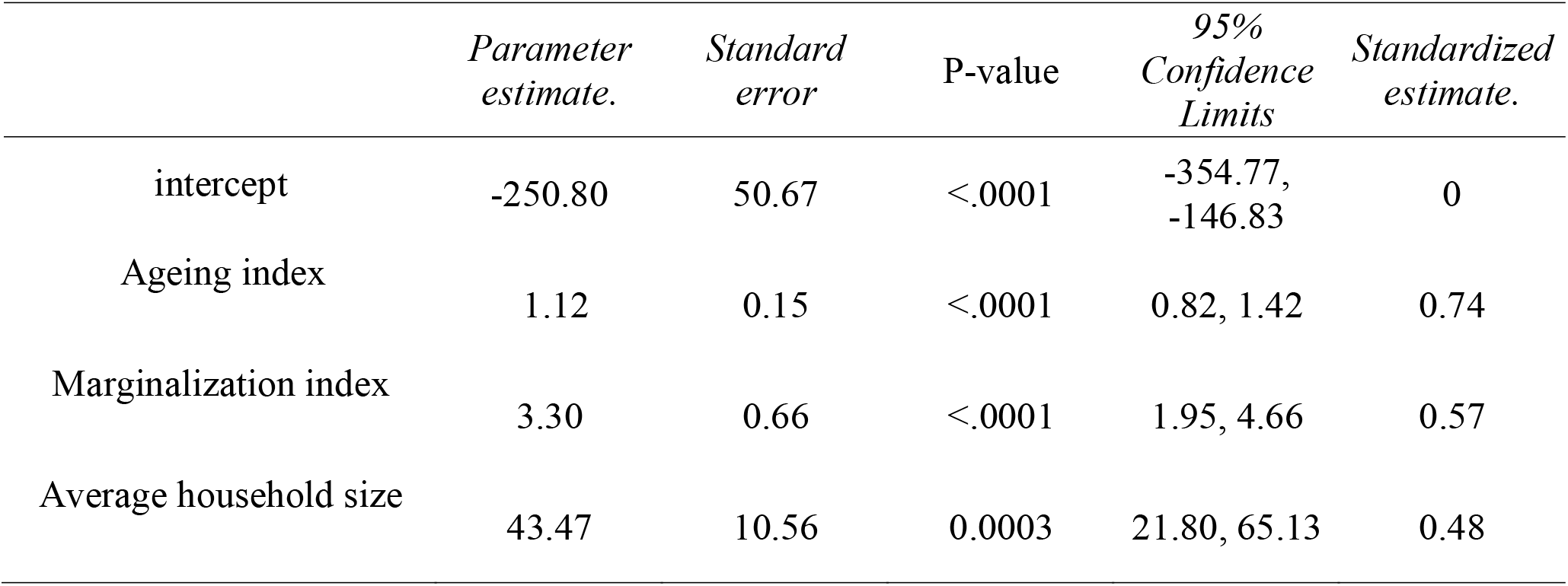
Results for the Final Regression Model 4 of excess mortality rate at the state level in Mexico (*n*=31)

|  | *Parameter estimate.* | *Standard error* | P-value | *95% Confidence Limits* | *Standardized estimate.* |
| --- | --- | --- | --- | --- | --- |
| intercept | -250.80 | 50.67 | <.0001 | -354.77, -146.83 | 0 |
| Ageing index | 1.12 | 0.15 | <.0001 | 0.82, 1.42 | 0.74 |
| Marginalization index | 3.30 | 0.66 | <.0001 | 1.95, 4.66 | 0.57 |
| Average household size | 43.47 | 10.56 | 0.0003 | 21.80, 65.13 | 0.48 |

The results of our clustering analyses is displayed in a dendrogram plot (Supplemental Figure 1). Specifically, we identified the following four prominent clusters based on the shapes of excess growth rate curves at state level:

*Cluster 1:* Baja California, Coahuila, Guanajuato, Hidalgo, Jalisco, Mexico, Mexico City, Michoacan, Morelos, Nayarit, Nuevo Leon, San Luis Potosi

*Cluster 2*: Aguascalientes, Chihuahua, Durango, Queretaro, Zacatecas

*Cluster 3*: Baja California Sur, Colima, Guerrero, Oaxaca, Puebla, Quintana Roo, Sinaloa, Sonora, Tabasco, Tamaulipas, Veracruz, Yucatan

*Cluster 4*: Campeche and Chiapas

**Supplemental Figure 1.**
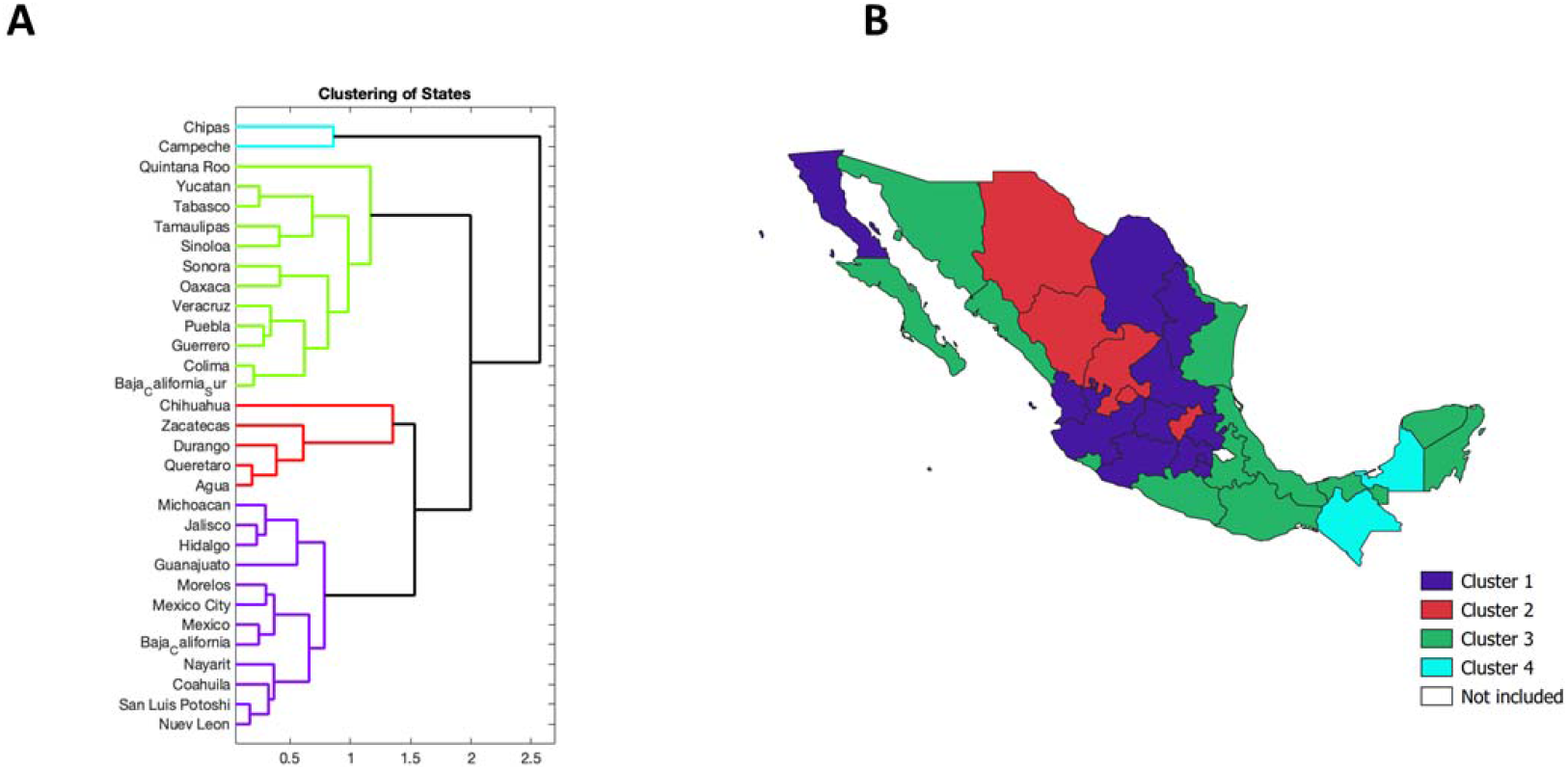
A: Dendrogram plot, B: map of Mexico showing the states in four different clusters

Figure 3 shows the average growth rate curves and one standard deviation band around it. The growth patterns in each cluster are very distinct. For cluster 1, we see two different peaks in growth rate, first small peak in July 2020 and the second big peak in January 2021. For cluster 2, there is a rapid increase in growth rate since July 2021 that peak on around December 2020. Unlike cluster 2, in cluster 3, the first big peak in July is followed by a small peak in January. Finally, in cluster 4, the growth rate rapidly increases from April to July followed by a rapid fall and a small rise in January 2021. In overall, first peak in most of the states occurred in around July, 2020 and the second peak occurred in around January, 2021.

**Figure 3.**
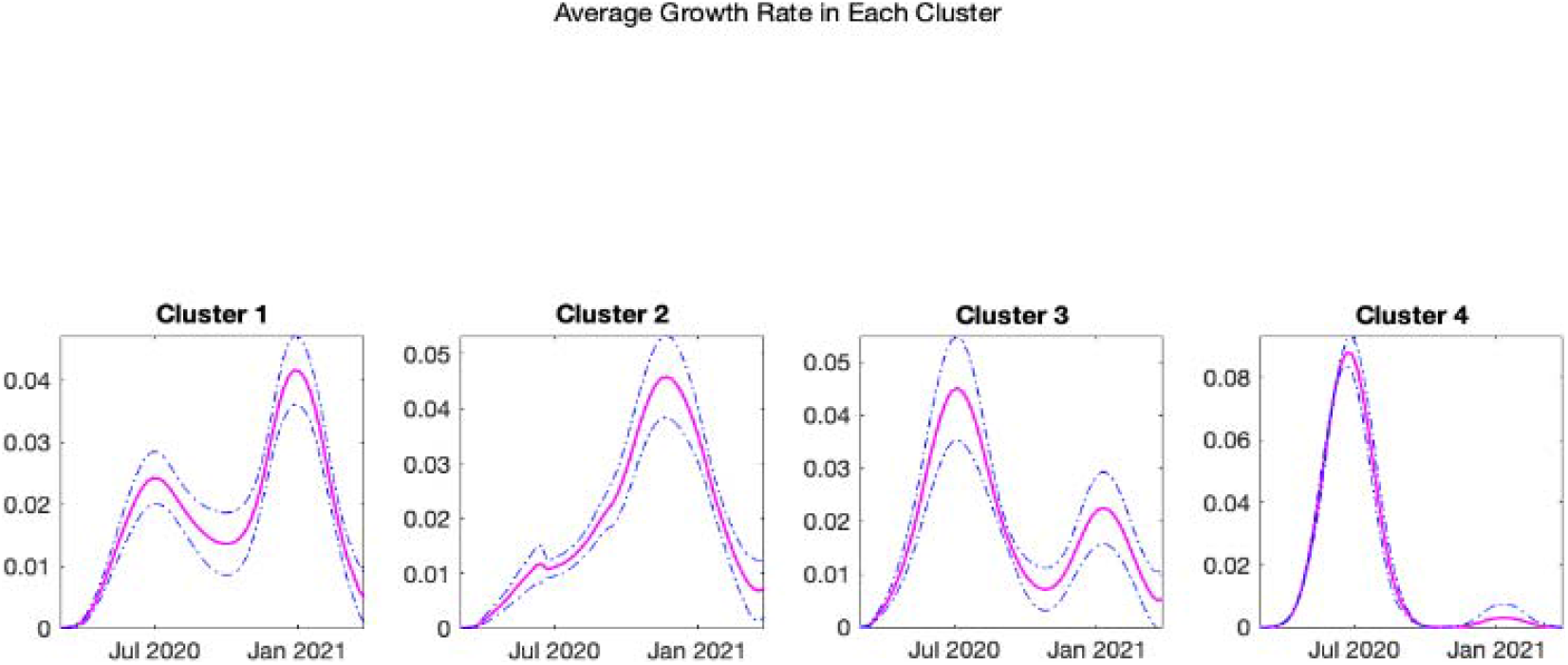
Average growth rate in each cluster, the dotted blue line are the one standard deviation band around the average growth rate

## Discussion

In this study we investigated the excess mortality patterns during COVID-19 pandemic at the national and subnational level in Mexico from March 1, 2020, to April 10, 2021. We estimated an excess all-cause mortality rate of 39.66 per 10 000 population at the national level (a total of ∼508 289 excess deaths), indicating a devastating mortality impact of COVID-19 pandemic in Mexico. We found that the excess mortality rate has continuously declined after the second COVID-19 peak during the week of January 17-23, 2021. Mexico City alone accounted for about 19% of total excess deaths in Mexico, with an excess mortality rate of 106.17 per 10 000 population.

Interestingly, we found that the states with the highest excess death rate (i.e., Mexico City, Tlaxcala, Morelos, Puebla, Mexico) were the central states in Mexico, while the lowest excess death rates were observed in the southern states (i.e., Chiapas, Oaxaca, Quintana Roo, Yucatan, Campeche). In Mexico, the majority of the indigenous population are located in the southern states (29). In 2015, Oaxaca had the highest proportion of native population (65.73%), followed by Yucatan (65.40%), Campeche (44.54%), and Quintana Roo (44.44%) (30). Compared to non-native groups, the indigenous populations across continents have suffered health disparities and a greater burden of diseases, including higher infant mortality, and lower life expectancy (31). During the pandemic, the indigenous populations have remained at higher risk of infection and death (32, 33). A previous study demonstrated higher excess deaths in U.S. states with higher concentration of native Americans (34). In contrast, in this study we found a lower excess mortality rate during the pandemic in Mexican states with a higher proportion of the native population. This is an intriguing finding that warrant further inquiry and examination as it may provide great insight to factors that may buffer against the impact of the pandemic and other adverse health events including natural disasters.

In our analyses of data form Mexico, we found that COVID-19 specifc deaths accounted for only 42.16% of total excess death at the national level, lowest in Chiapas (20.97%) and highest in Quintana Roo (76.05%). At the state level in Mexico, the timing and the rigor of implementation of public policies to contain the virus has varied widely (16). For example, some of the states such as Veracruz, Yucatan, Nuevo Leon, and Tamaulipas, established policies to promote social distancing before the federal government enacted those policies (16). While states such as Chiapas, Tabasco, San Luis Potoshi, and Zacatecas underperformed in implementation of public policy measures (16), some other states, with a relatively low excess mortality rate, such as Baja California Sur and Nayarit implemented public information campaigns and international travel restrictions for longer periods, despite the potential adverse impact on tourism, which is a major economic activity (16).

We found a positive association between the aging index and excess mortality, in the adjusted model confirming previous studies linking older age and COVID-19. The aging index is defined as the number of older adults (60 years of age and older) for every 100 children and youth (0 to 14 years of age)(35), and it increases as the population ages. Older age is a significant predictor of COVID-19 mortality as well as mortality from other causes(36) (37) (38). Previous studies that analysed excess mortality pattern during the first wave of COVID-19 in 21 industrialized countries has shown that those aged 65 years and above comprised 94% of all excess deaths, indicating a very high risk of death among older aged population specifically due to COVID-19 (15).

Similarly, our finding of a positive association between the marginalization index and and excess mortality support previous research underscoring the close link between social disadvantage and COVID mortality and the overall increased burden of the pandemic in marginalized populations. The marginalization index that we used is an indicator of the inequities in quality of housing, access to basic public services like electricity and drinking water, schooling, proportion of poorly paid population and other sociodemographic and population health characteristics (21, 39). There may be several explanations for our findings. For example, public health measures such as social distancing and sheltering in place to combat the COVID-19 pandemic resulted in a disproportionate burden to vulnerable and marginalized populations (40-42). Marginalized groups are also more likely to be infected by the coronavirus due to their the context of their living arrangements and which may limit the ability to self-isolate and socially distance. Similarly, it is well demonstrated that marginalized populations tend to have a higher prevalence of chronic conditions such as obesity, hypertension and diabetes which are all strong risk factors associated with poor prognostic outcomes among those infected with COVID-19 (42). To complicate matters further, marginalized population are also often at greater risk of dying due to other indirect causes such as limited access to already-stressed health care system, poor mental health outcomes, food insecurity, abuse and violence among other social ills. (41, 42).

Interestingly, Chiapas, a Southern Mexico with a high marginalization index(21), a higher concentration of indigenous population(30), and lower average performance in implementation of public policies to combat COVID-19 (16), had the lowest excess mortality rate among the states examined in these analyses. While there is no clear explanation for these finding, the varying climate across the states examined may be a contributing factor. For example, the southern states in Mexico have a weak seasonality and a tropical climate throughout the year (43). According to previous studies conducted in Mexico, tropical climate delayed the local transmission of SARS-CoV-2 at regional level. As such, the temperate climate regions like Tlaxcala and Jalisco may have been more vulnerable for local transmission than the tropical climate regions such as Chiapas and Veracruz. These findings should be replicated in other setitngs that comprise multiple climate regions to determine the impact of seasonality in the transmission spread and impact of excess mortality patterns. Additionally, more research is needed to elucidate the factors associated with lower all-cause excess death rate in the relatively marginalized southern states as observed in this study. Such insight may provide mitigation strategies for other regions with higher impact.

We also found a positive association between average household size and excess death rate in the adjusted model. Although the links between average family size and excess death rate at the state level have not been reported elsewhere, the average family size could have interacted with other social determinants of health such as poverty, food insecurity, and lack of access to health care. Further studies are needed to understand the potential mechanism underlying this association and to more specifically consider family size as a potential population-level indicator of communities at risk for increased impact.

Our classification of excess deaths growth rate curves at the state level reflects four distinct categories of Mexican states. In all of the clusters the first peak of excess deaths growth rate curve occurred in around July, 2020 which happened after the phased reopening of non-essential services in June, 2020 in Mexico. The reopening of the country coincided with both increase in driving and walking trends, and the highest levels of COVID-19 deaths that remained at a high level during June and July 2020 (44). The visual analysis of the growth rate curve indicates that the coastal tropical southeastern states were most affected during the first few months of the pandemic compared to other states. However, these states exhibited the lower overall excess death rate that could indicate the effect of temperature and other environmental factors. Moreover information on the growth rate curves can be utilized at the state level to guide the implementation of medical and public health measures. Besides, learning from the public health measures implemented in states of one cluster (for example, cluster 4) can be helpful to the other states (for example, states in cluster 1).

To our knowledge, this is the first study that assesses the growth rate curves of excess deaths at the state level in Mexico. In our study, the estimates of excess death rate, as well as the proportion of COVID-19 attributed deaths, could be underestimated due to factors such as low testing rates in Mexico, misclassification of COVID-19 deaths, and delay in reporting COVID-19 deaths. This limitation should be considered when interpreting these findings. Additionally, other potential confounders that were not measures may explain the patterns of excess mortality across states.

## Conclusion

Our estimate of all-cause excess death rate in Mexico was 39.66 per 10 000 with central states exhibiting higher rates and southern states exhibiting lower rates. Our study highlights that several population measures including the aging index, marginalization index, and average household size were significantly associated with the all-cause excess mortality rates across Mexican states during the COVID-19 pandemic. Our excess mortality estimates can help tailor state specific medical and public health interventions to prevent excess mortality in vulnerable areas but targeting specific regions and socio-economic indicators. We also recommend further studies that investigate the lower excess death rate in southern states, and studies that explore the role of environmental factors, particularly the social determinants of health, in spatial variation in excess death rate in Mexico and other regions heavily impacted by COVID-19.

## Data Availability

All the data used in the study are publicly available. The sources of those data are provided under the methods section.

## Declarations

### Ethics approval

Not applicable

### Author contribution

GC and SD conceptualized and designed the Study. GC directed the study’s implementation. SD conducted the formal analysis and drafted the manuscript. RL, MS, GC reviewed and edited the manuscript. All the authors have read and approved the final version of this manuscript.

### Data availability

All the data used in the study are publicly available. The source of those data are provided under the methds section.

